# The Learning Early Infant Feeding Cues (LEIFc) Intervention to Increase Responsive Infant Feeding: A Proof-of-Concept Study

**DOI:** 10.1101/2025.03.26.25324053

**Authors:** Jessica Bahorski, Julie May McDougal, Edie Kiratzis, Connie Burke, Jalina Hess, Mollie Romano

## Abstract

**Background:** Interventions are needed to promote responsive infant feeding (RIF) to assist caregivers recognize infant hunger/satiety cues and overcome barriers to using RIF. The Learning Early Infant Feeding Cues (LEIFc) intervention was designed to fill this gap by using a validated coaching approach to promote RIF. Guided by the ORBIT model, this multimodal, proof-of-concept (POC) study evaluated the clinical significance, feasibility, and fidelity of LEIFc in mother-infant dyads.

**Methods:** Mother-infant dyads were followed from the 3rd trimester of pregnancy to 4 months postpartum. Coaching during a feeding session was provided using the SS-OO-PP-RR (“super,” Setting the Stage, Observation & Opportunities, Problem Solving & Planning, Reflection & Review). RIF was measured pre-post via the *Infant Feeding Questionnaire* subscales: awareness of infant cues (Aware), feeding on a schedule (Schedule), and using food to calm (Calm). Qualitative data from participants was collected at study conclusion.

**Results:** 21 dayds completed all study visits. Suggesting RIF, there was a 28.5% increase in Aware and a 40% decrease in Calm. Schedule increased 4.8%, not suggestive of RIF. Three themes emerged from the qualitative data: LEIFc offered additional support, recognition of infant feeding cues, and identification of additional needs

**Conclusions:** This POC study supports that the LEIFc intervention was feasible to implement and acceptable by participants. LEIFc was effective at increasing RIF except feeding on a schedule. The study concluded at 4 months postpartum when many mothers return to work and/or have other caregivers feeding their infant which may contribute to feeding on a schedule. Mixed methods analyses may explain the results. Consistent with the ORBIT model, the next steps are to refine the intervention then test in a larger sample at highest risk for poor infant outcomes associated with feeding (i.e., rapid infant weight gain, childhood obesity).

---

Responsive feeding is important for infant self-regulation of food consumption (Hodges et al., 2016; Richardson, Ventura, et al., 2024). Responsive feeding occurs when the caregiver recognizes an infant’s cues of hunger and fullness then responds promptly to these cues (Hodges et al., 2016; Pérez-Escamilla et al., 2017). By feeding in response to cues, opposed to premeasured amounts of breastmilk or formula or a strict schedule, overfeeding is prevented. Preventing overfeeding is important for maintaining proper weight to height ratios across infancy which has been shown to prevent future childhood obesity. The first weeks after birth are critical for establishing infant feeding practices (Paul et al., 2011; Radzyminski & Callister, 2016; Thompson & Bentley, 2013). Education on and promotion of responsive feeding should begin prenatally and evolve with the development of the child (Pérez-Escamilla et al., 2017). The responsive feeding approach can be used with milk-based feedings, from the breast or bottle, and continued when complementary or table foods are introduced. This provides tremendous value across development stages and cultural feeding practices and beliefs. For this reason, the responsive infant feeding approach is recommended by organizations such as the American Academy of Pediatrics (AAP) and American Speech-Language-Hearing Association (ASHA), and is included in the Dietary Guidelines for Americans (AAP Committee on Nutrition, 2025; United States [U.S.] Department of Agriculture, 2020; ASHA, n.d.). However, parents and other caregivers do not always understand how to recognize and respond to infant feeding cues, thus education is needed to determine how to incorporate responsive feeding into infant feeding practices (Redsell et al., 2021; Richardson, Ventura, et al., 2024).

A systematic review found that enablers to responsive infant feeding included recognizing infant cues of hunger and satiety, infant feeding knowledge, and family/friend support (Redsell et al., 2021). Additionally, mothers who breastfeed their infant tend to be more responsive in their feeding approach (Ventura, 2017). However, there are barriers to use of responsive infant feeding such as parental desire for control over infant feeding and healthcare professional advice regarding breast-or formula feeding (Redsell et al., 2021). A study of Special Supplemental Nutrition Program for Women, Infants, and Children (WIC) participants and counselors found that education and training on responsive infant feeding specific to bottle feeding is needed (Richardson, Ventura, et al., 2024). Culture beliefs and practices are also important to consider as it is well known that culture plays a large role in eating preferences and behaviors (de Diego-Cordero et al., 2021; Hamner et al., 2021; Kim et al., 2017; Roll & Cheater, 2016). Maternal mental health, such as high levels of stress, depression, or anxiety, have been associated with a non-responsive feeding approach (Hurley et al., 2008; Richardson, Reis, et al., 2024; Thompson et al., 2021). Future interventions should consider these constructs.

Large randomized controlled trials (RCTs), Intervention Nurses Start Infants Growing on Healthy Trajectories (INSIGHT), NOURISH, and Mothers and Others Study, have included a responsive feeding component within their intervention (Daniels et al., 2009; Paul et al., 2014; Wasser et al., 2017). All studies showed significant, positive, differences between intervention and control group in regards to responsive feeding and/or dietary behaviors (Daniels et al., 2013; Daniels et al., 2015; Hohman et al., 2017; Savage et al., 2018; Thompson et al., 2021). The INSIGHT study also showed that intervention infants gained weight more slowly than control infants regardless of feeding mode suggesting that rapid weight gain was prevented (Savage et al., 2016). The NOURISH and Mothers and Others studies showed a trend toward slower weight gain that did not reach statistical significance (Daniels et al., 2013; Daniels et al., 2015; Wasser et al., 2020). It should be noted that differences existed in these studies which may have contributed to the varied results. INSIGHT and NOURISH participants were primarily high-income, married, well-educated, and non-Hispanic White (Daniels et al., 2009; Daniels et al., 2013; Paul et al., 2014; Savage et al., 2016), whereas Mothers and Others participants were non-Hispanic Black participants, mostly single and low income (Thompson et al., 2021; Wasser et al., 2020; Wasser et al., 2017). Only the Mothers and Others study began with a prenatal visit when maternal decisions regarding infant feeding usually begin; the first visit for the other two studies was postnatally. Additionally, all measurement of responsive infant feeding was by maternal self-report (Daniels et al., 2009; Paul et al., 2014; Wasser et al., 2017); an objective measurement of responsive feeding may provide a different perspective of responsive feeding by picking up infant cues and maternal response to these cues. Finally, to increase sustainability, more research is needed to test responsive feeding interventions within existing programs, beyond a research protocol.

Outcomes of these studies and gaps should be considered in future intervention development focused on responsive infant feeding. There is a need for research on a sample that is diverse in income, marital status, education level, and race/ethnicity to compare differences across demographic groups. If interventions are focused on a specific cultural group, then materials should be developed that are most suitable and effective for that group (Hernandez et al., 2022; Richardson, Ventura, et al., 2024; Rosenstock et al., 2021).

Measurement of responsive feeding should be considered, as maternal self-report versus an objective measure can provide different outcomes; objective measures of responsive feeding do exist (Hodges et al., 2013; Sumner & Spietz, 1994). Finally, measurement of change in use of the responsive feeding approach pre-post intervention and across time points should also be considered.

To fill some of these gaps, the Learning Early Infant Feeding Cues (LEIFc) intervention was developed (Bahorski et al., 2023). The goal is to collaborate with maternal-child home visiting programs to implement the intervention to promote sustainability within existing programs aimed to improve maternal-child outcomes. The purpose of this study was to test proof-of-concept of the LEIFc intervention in a group of mothers from community settings representative of the area. A multimodal, quasi-experimental study design was used to examine the feasibility and social acceptability of the intervention. Findings will be used in the next step to refine the intervention and then test with mother-infant dyads enrolled in maternal-child home visiting programs using home visiting personnel as interventionists.

## Methods

The Obesity-Related Behavioral Intervention Trials (ORBIT) framework is used to guide development of this behavioral intervention (Czajkowski et al., 2015; Powell, 2021). Methods for this study were previously described (Bahorski et al., 2023). Data collection occurred from May 2022 to October 2023 (recruitment and enrollment May 2022 to April 2023). The study was approved by the Florida State University, Institutional Review Board.

### Participants

Mothers were recruited in the 3^rd^ trimester of pregnancy (28 weeks or beyond) from community settings. Eligible participants were pregnant with a singleton infant with no known congenital or genetic anomalies, able to read and understand English or Spanish, and 18 years of age or older. Participants were recruited from obstetric offices, social media groups that target pregnant mothers, and word of mouth. Mothers were consented upon enrollment in the study; dyads were rescreened for eligibility after birth of the infant and the mother provided consent for her infant if still eligible. Due to being a minor risk study, consent from the father of the infant was waived.

### Study Procedures

Study Visit 1 occurred between 28 weeks and the birth of the infant and was conducted via the video conferencing platform Zoom. Study Visits 2 to 5 occurred following birth at infant age of 1, 2, 3, and 4 months respectively. The majority of visits were conducted in participants’ homes, however, the option for a clinic visit was provided for participants who were not comfortable with study visits in their home. Infant length and weight were collected at Study Visits 2-5 as well as infant feeding practices (i.e., breastmilk [from breast and/or bottle], formula, introduction of complementary foods). Study procedures are outlined in Table 1.

**Table 1.** LEIFc Study Visits.

| **Table 1. LEIFc Study Visits** |  |  |  |  |
| --- | --- | --- | --- | --- |
| Visit | Timing | Components | Survey | Compensation (all via gift cards) |
| 1 | Prenatal | • Video demonstrating infant cues of hunger and satiety<br>• Brochure on infant feeding<br>• Zoom session with interventionist to discuss cues and answer questions | • Prenatal Questionnaire Demographics<br>• MIBS<br>• PHQ-9 | \$15 |
| 2 | 1 month PP (pre-intervention) | • Video of feeding session no coaching, coded for responsive feeding | • Birth History<br>• MIBS<br>• PHQ-9<br>• IFQ | \$25 |
| 3 | 2 months PP | • Video of feeding session with COACHING, measure of fidelity | • MIBS<br>• PHQ-9 | \$25 |
| 4 | 3 months PP | • Video of feeding session with COACHING, measure of fidelity | • MIBS<br>• PHQ-9 | \$25 |
| 5 | 4 months PP (post-intervention) | • Video of feeding session no coaching, coded for responsive feeding | • MIBS<br>• PHQ-9<br>• IFQ | \$35 |
| Note. MIBS = Mother-Infant Bonding Scale; PHQ-9 = Patient Health Questionnaire-9; IFQ = Infant Feeding Questionnaire; PP = postpartum |  |  |  |  |

### Intervention

The LEIFc intervention was developed by the research team and includes several key components, including a prenatal visit to introduce infants’ early feeding cues and in the moment coaching supports during feeding routines after the baby is born. The SS-OO-PP-RR (“super,” Setting the Stage, Observation and Opportunities, Problem Solving and Planning, Reflection and Review) coaching approach is a hallmark of the intervention that is intended to build caregiver confidence in reading their child’s cues with in the moment practice and feedback as well as problem solving and reflecting on what is working or what needs to be adapted.

Originally developed to support caregivers of infants and toddlers with disabilities to embed intervention in daily routines (Windsor et al., 2019; Woods, 2021), SS-OO-PP-RR was adapted to be specific to infant feeding and maternal communication with an infant during the first few months of life. SS-OO-PP-RR is characterized by a partnership between the interventionist and caregiver (mothers in this study) to identify components of infant feeding specific to responsive feeding and implement responsiveness strategies to increase recognition of infant feeding cues and how to respond promptly to these cues. SS-OO-PP-RR is unique from other responsive feeding interventions’ coaching approaches by individualizing the techniques to each family, offering opportunities to practice, reflect, problem solve and plan to help each caregiver interpret and respond to their child’s cues.

### Measurement tools

#### Quantitative data

The Infant Feeding Questionnaire (IFQ) is a 20-item, self-report measure of maternal beliefs and practices regarding infant feeding, with seven subscales: Concern for Underweight (4 items), Concern for Hunger (3 items), Awareness of Infant Cues (4 items), Concern for Overweight (3 items), Feeding on a Schedule (2 items), Use of Food to Calm (2 items), and Social Interaction (2 items). All responses are on a 5-point, Likert scale; higher scores indicate a stronger measure of each construct. Three of the subscales suggest responsive feeding: an increase in awareness of infant cues, and a decrease in feeding on a schedule and providing food to calm an infant. Awareness of cues is measured by questions such as “I knew when he/she was hungry” and “I knew when he/she was full” with responses ranging from “disagree a lot” to “agree a lot.” Feeding on a schedule is measured with questions “Did you let him/her eat whenever he/she wanted to?” and “Did you only allow him/her to eat at set times?” Responses ranged from “never” to “always;” “Did you let him/her eat when he/she wanted to?” was reversed scored. Using food to calm is measured with questions, “Feeding him/her was the best way to stop his/her fussiness,” with responses ranging from “disagree a lot” to “agree a lot,” and “When he/she got fussy/was feeding him/her the first thing you would do?” with responses ranging from never to always.

The Patient Health Quesitonnaire-9 (PHQ-9) is a well validated tool to screen for depression. The 9-item self-report survey asks individuals to rate their feelings over the past 2 weeks with answers of “not at all,” “several days,” “more than half the days,” and “nearly every day.” Scores range from 0 to 3 for each question providing a total score range from 0-27 with higher scores suggesting greater depression severity.

The Mother-Infant Bonding Scale (MIBS) is an 8-item measure of mother-infant bonding. Mothers are asked to rate their feelings toward their infant in the first weeks after birth from the following adjectives: loving, resentful, neutral or felt nothing, joyful, dislike, protective, disappointed, and aggressive. Likert-type responses range from “very much” to “not at all” with scores ranging from 0 to 3 for each response and total scores of 0-24; higher total scores indicate greater difficulty in bonding.

Video recordings of each feeding session (Visits 2-5) were completed with iPads and a tripod. The recordings at Visit 2 and 5 were observations of mother-infant interactions and served as an objective measurement of responsive feeding pre-post intervention. These data are being analyzed and will be presented at a later date. Visits 3 and 4 were coaching intervention sessions using the SS-OO-PP-RR framework. A random selection of 30% of coaching sessions were scored for implementation fidelity (Appendix A) by a trained research assistant who was not involved in the intervention.

#### Qualitative data

After Visit 5, the final study visit, participants were asked open ended questions regarding their experience participating in the research study and receiving the intervention; along with questions regarding infant feeding (Table 2). This visit was conducted by research assistants and not the interventionist. The session was audio recorded using an iPad, no video recording was done during this portion of the study visit.

**Table 2.**
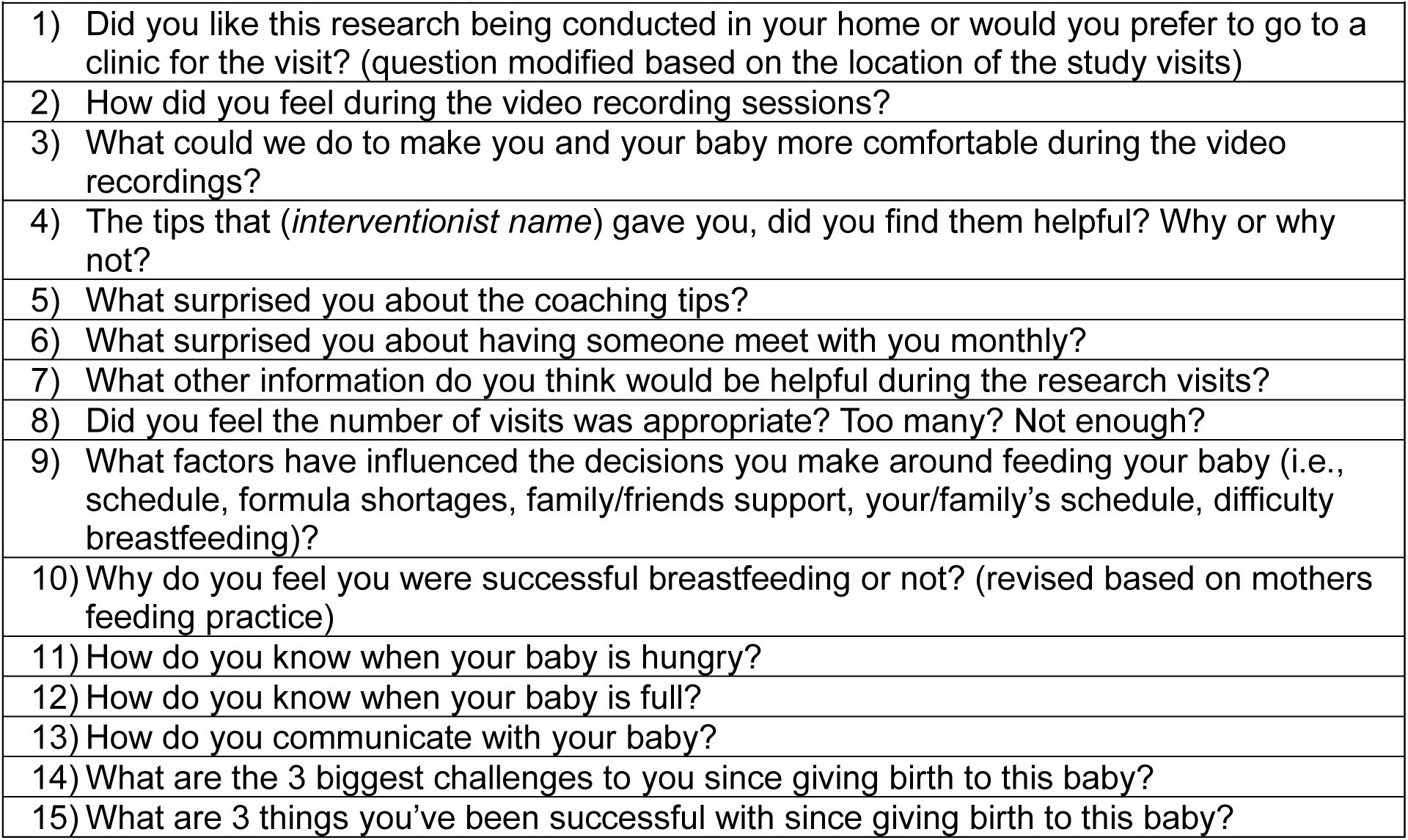
Qualitative research questions.

|  |
| --- |
| Table 2. Qualitative research questions |
| 1) Did you like this research being conducted in your home or would you prefer to go to a clinic for the visit? (question modified based on the location of the study visits) |
| 2) How did you feel during the video recording sessions? |
| 3) What could we do to make you and your baby more comfortable during the video recordings? |
| 4) The tips that (*interventionist name*) gave you, did you find them helpful? Why or why not? |
| 5) What surprised you about the coaching tips? |
| 6) What surprised you about having someone meet with you monthly? |
| 7) What other information do you think would be helpful during the research visits? |
| 8) Did you feel the number of visits was appropriate? Too many? Not enough? |
| 9) What factors have influenced the decisions you make around feeding your baby (i.e., schedule, formula shortages, family/friends support, your/family's schedule, difficulty breastfeeding)? |
| 10) Why do you feel you were successful breastfeeding or not? (revised based on mothers feeding practice) |
| 11) How do you know when your baby is hungry? |
| 12) How do you know when your baby is full? |
| 13) How do you communicate with your baby? |
| 14) What are the 3 biggest challenges to you since giving birth to this baby? |
| 15) What are 3 things you've been successful with since giving birth to this baby? |

### Data analysis

#### Quantitative data

Descriptive statistics were analyzed on all participants who completed Visit 1. Due to the small sample size, variables were condensed to eliminate excessive demographic categories. For example, race was divided into White, Black, and other/mixed races. Education was divided into those who had completed a college degree and those who had not. The PHQ-9 and MIBS were totaled as appropriate to the tool. As a proof-of-concept study, it is important to understand factors that may have contributed to study drop out thus, demographic characteristics were compared between the participants who completed the study and those who did not. Fisher’s Exact test or Mann-Whitney U test were used as appropriate to the variable.

To examine clinical significance, changes in three subscales of the IFQ were examined: Awareness of Infant Cues, Feeding on a Schedule, and Use of Food to Calm. A score of 4 or higher on the IFQ indicates that the practice was used “often” or “always;” thus examining the percent of mothers who reported a score of 4 or higher pre- and post-intervention was examined and the difference calculated.

Statistical significance was also examined. Each of the IFQ subscales pre-post intervention was totaled and normality assessed. The assumptions of normality were met for the use of food to calm variable but not for the remaining variables. Thus, paired t test was used to examine use of food to calm pre-post intervention, and Wilcoxon signed rank test was used for the other variables. Effect sizes were calculated to determine the magnitude of effect. Due to the small sample size, covariates were not considered in analyses.

#### Qualitative data

De-identified audio recordings were transcribed by Ubiqus On Demand (https://www.ubiqus.io/) and then uploaded to NVivo software version 14 for analysis. Research assistant (CB) reviewed all transcripts and decided on 16 initial codes. Principal investigator (JB) then reviewed the transcripts and codes and the two members of the research team met to ensure consistency in codes and remove any discrepancies. From there, thematic analysis occurred and two sets of themes emerged; one specific to the intervention and research methods, and a second specific to feeding practices. The recordings, transcripts, codebook, and final themes provide an audio trail to ensure transparency. For purposes of this manuscript, only the themes specific to the intervention and research methods are presented.

## Results

Twenty-nine (29) mothers consented to participate in the study with 21 completing all study visits. One participant had her baby before Visit 1 (prenatal visit) could be completed, the remaining 7 dropped out or were lost to follow up between study Visit 1 and study Visit 2 (1 month postpartum) (Figure 1). We examined the differences in participants who completed the study versus those who did not (Table 3). No significant differences were seen between groups, but three areas trended toward significance; a larger percentage of participants who identified as Hispanic did not complete the study (57.1% vs 9.5%, *p* = 0.09) and more mothers who completed the study planned to exclusively breastfeed their infant (95.2% vs 71.4%, *p* = 0.14) and reported a higher score on the prenatal depression screen (5.9 vs 4.0, *p* = 0.11).

**Figure 1.**
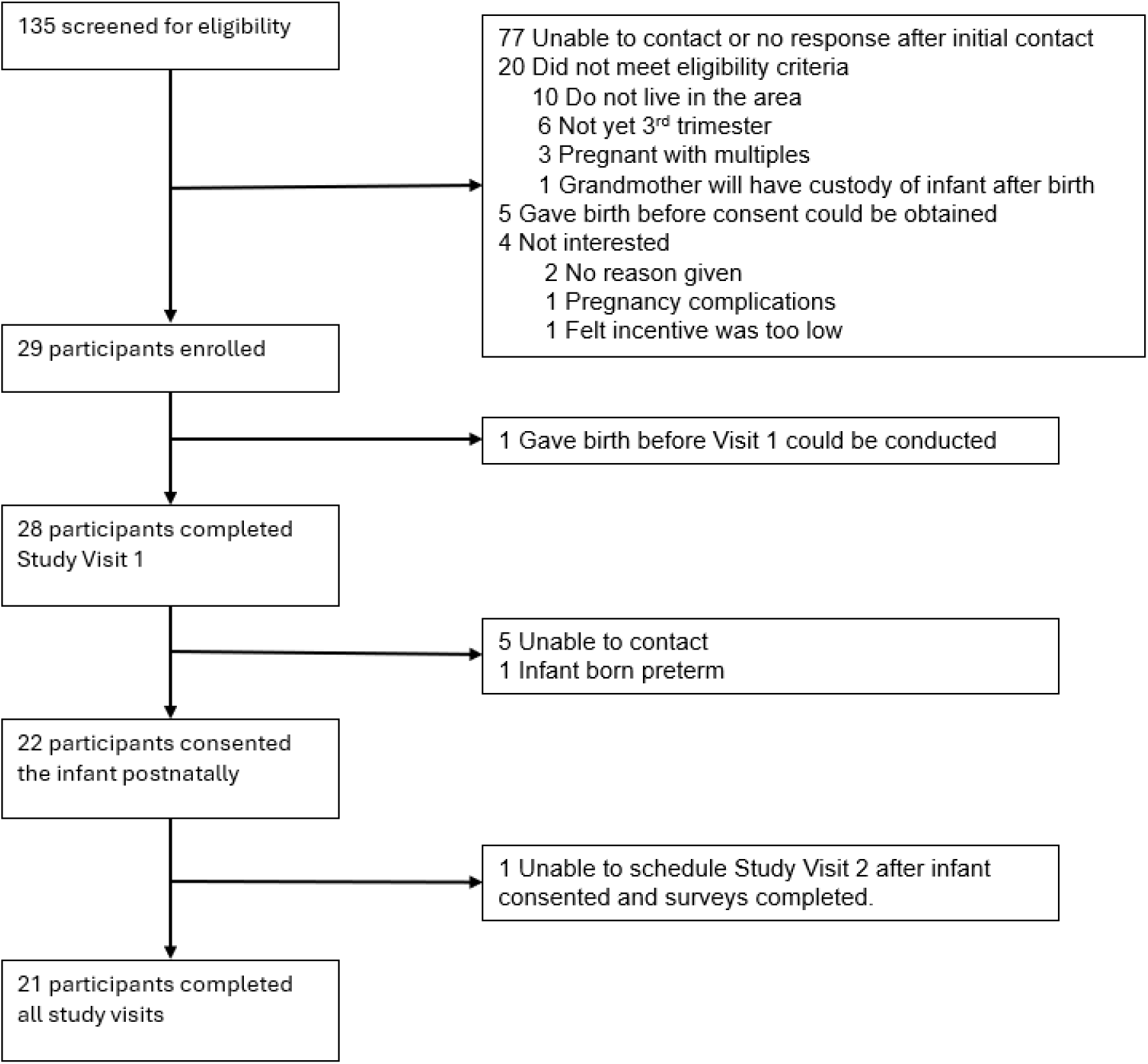
Flow diagram.

**Table 3.**
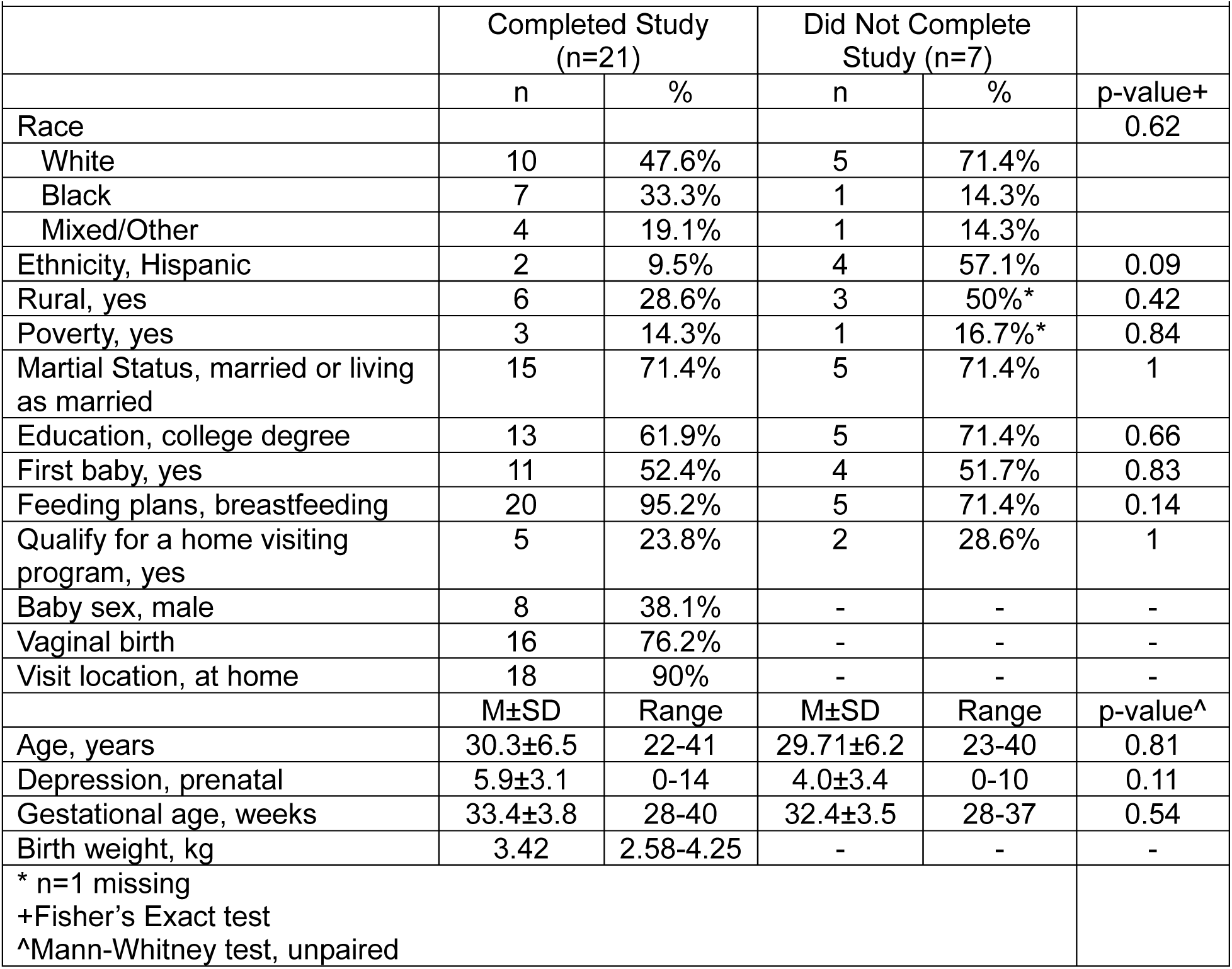
Comparison of those who completed the study vs those who did not (N=28).

Participants who completed all study visits were primarily married or living as married (71.4%), held a college degree (61.9%), and were working full-time (52.4%). The average age of mothers was 30.3 years, and the sample was racially and ethnically representative of north-central Florida. Ninety percent (90%) of mothers had all visits completed in their home with the remaining occurring at the research clinic.

Thirty-one percent (31%) of intervention sessions were coded for fidelity using the implementation checklist (Appendix A). Mean fidelity was 82 with a range of 56.25 to 100. The low end of the range was attributed to a fussy, unhappy infant making coaching difficult during those times. The mean of 82 is consistent with prior work using the SS-OO-PP-RR coaching approach (Romano et al., 2021; Romano et al., 2023).

### Quantitative Data

The primary outcome of this study was use of responsive feeding pre-post intervention. As a proof-of-concept study, clinical significance was examined. Suggestive of responsive feeding, there was a 28.5% increase in awareness of cues and a 40% decrease in using food to calm of participants reporting a score of 4 or higher. Feeding on a schedule also increased 4.8% which is not consistent with responsive feeding (Figure 2).

**Figure 2.**
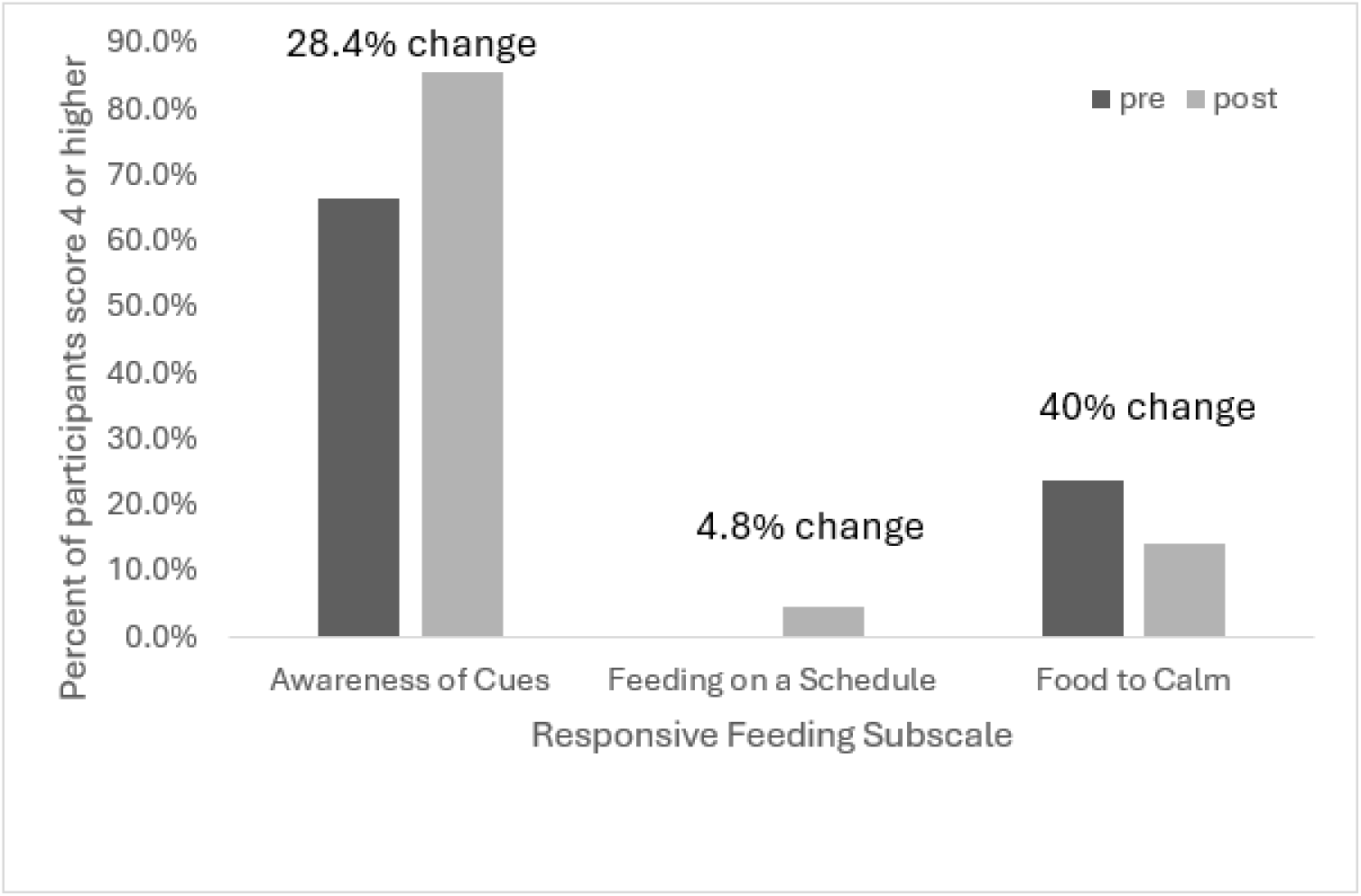
Measure of clinical significance.

There was a statistically significant, medium increase in awareness of cues (M = 4.167 vs M = 4.381, *p* = 0.05, *r* = 0.43) and in feeding on a schedule (M = 1.381 vs M = 1.643, *p* = 0.05, *r* = 0.45). There was a small decrease in using food to calm which did not reach significance (M = 3.000 vs M = 2.786, *p* = 0.12, *d* = 0.23). An increase of awareness of cues and a decrease in feeding to calm suggest use of responsive feeding. The increase in feeding on a schedule does not suggest use of responsive feeding. The concern for hunger was the only other subscale on the IFQ that trended toward significance showing a medium increase from pre-to post-intervention (M = 1.03 vs M = 1.22, *p* = 0.07, *r* = 0.41) (Table 4).

**Table 4.**
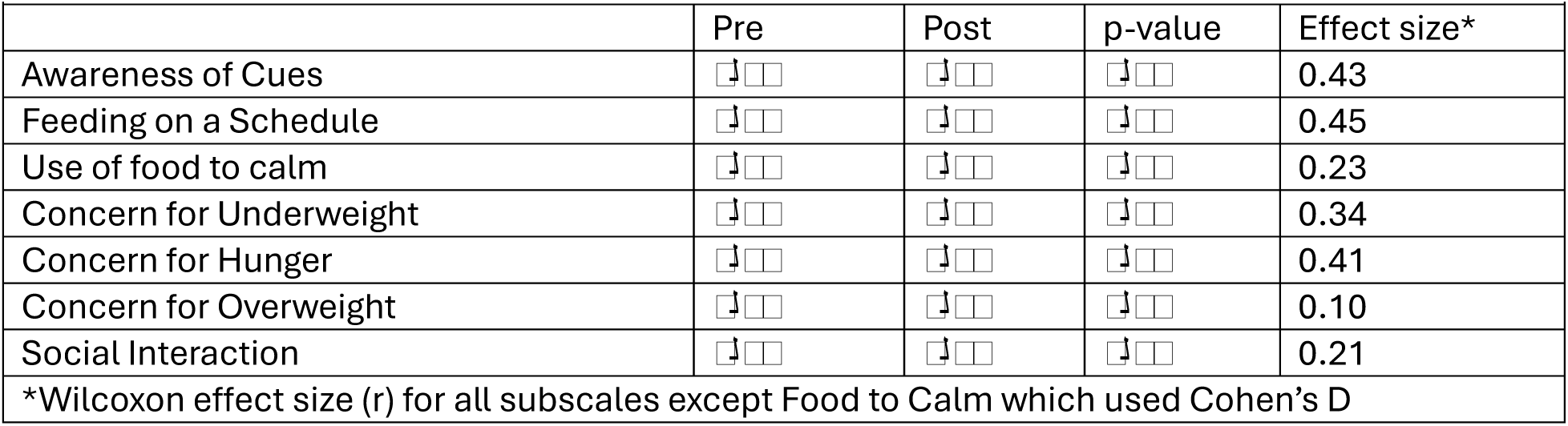
IFQ Subscale scores pre-post intervention (N=21).

### Qualitative Data

Qualitative data collected from mothers at study conclusion indicated that the study was feasible and acceptable by participants. Three themes emerged specific to the LEIFc intervention: 1) LEIFc offered additional support; 2) Recognition of infant cues; and 3) Additional needs.

### LEIFc offered additional support

Participants expressed that they really enjoyed having the interventionist come to their home. They liked the relationship developed with the interventionist and felt that having someone to brainstorm solutions/ideas and offer reassurance was beneficial. The participants also felt validated in their parenting and feeding decisions.

*“I thought it was really great to have someone consistently come out and just assess and check in…”*

*“She was basically just validating every baby is different and you don’t have to do that and it’s okay to do your own thing.”*

*“I appreciated the rapport over time [and having] that touchpoint and checkpoint. I think I appreciated seeing similar researchers over and over…I thought that was good to build up a bit of a relationship.”*

*“I have found that the tips you gave me I could use in other aspects of parenting. I was more responsive to other interactions with my children.”*

### Recognition of Infant Feeding Cues

Consistent with the primary outcome of the intervention, to increase responsive infant feeding, the qualitative data supported that participants were aware of their infant’s cues.

Participants expressed not waiting for the infant to cry to feed him/her, not going straight to a bottle when the infant is crying, and that other needs may need to be addressed. The participants also expressed that they were able to better understand their infant.

*“Because I used to just wait for them to cry and tell me, so I thought that was their cue, but they have other cues.”*

*“With the feeding, it made me more aware to not just always go straight to a bottle every time he’s crying.”*

*“Some of the tips made me more conscious of what to look out for.”*

### Additional Needs

These data also provided insight regarding how to refine the intervention moving forward. Participants expressed the desire to learn more about developmental milestones and growth patterns of their infant. The need to extend the intervention through the introduction of complementary foods (i.e., solid foods) was strong. Complementary foods are often started between age 4 and 6 months; the intervention ended at 4 months right before this transition occurred. Finally, there was a lot of need for lactation support and topics related to bottle nipples sizes and breast pump flange sizes.

*“…like there’s some resources here if you need, your lactation questions or whatever, then that’s a helpful thing if you’re talking about going forward for other studies.”*

*“Maybe just more information about just different developmental milestones, and just how to reach them if you’re not reaching them.”*

*“…but just maybe tips for when we do start introducing food or other liquids besides just breast milk or formulas, might be helpful just for future.”*

## Discussion

This multi-modal, proof-of-concept study aimed to test the LEIFc intervention on the use of responsive feeding in mothers from community settings and determine feasibility and social acceptability. The study enrolled mothers during the 3^rd^ trimester of pregnancy and followed mother-infant dyads to 4 months postpartum. Unique to this intervention is the individualized SS-OO-PP-RR coaching approach to assist mothers in learning infant feeding cues and responding appropriately to these cues, which also increases overall communication between the mother and infant. The intervention was feasible to implement in family homes and was acceptable to the mothers who participated. Suggestive of the responsive feeding approach, participants’ self-report of awareness of their infant’s cues significantly increased pre-post intervention. Using food to calm their infant decreased which also suggests participants used the responsive feeding approach. Feeding the infant on a schedule increased pre-post intervention which suggests a non-responsive feeding approach. Results of this study also provided insight into how to refine the intervention moving forward, specifically, extending the intervention through the introduction of complementary foods, providing education on growth and development, and including more education and support specific to breastfeeding and pumping breastmilk.

As a proof-of-concept study, it is important to evaluate clinical significance opposed to statistical significance and to understand the feasibility and acceptability of the intervention (Czajkowski et al., 2015; Powell, 2021; Powell et al., 2018). The 28.4% increase in participants’ awareness of cues and 40% decrease in using food to calm demonstrate clinical significance of the intervention and warrant evaluating the intervention for refinement and moving through the next phases of the ORBIT framework. Our rate of enrollment, approximately 2.6 participants each month (29 participants over 11 months), needed to be higher to complete the study in 1 year as planned but was within the target range of 27-36 participants (Bahorski et al., 2023). We have identified ways to increase recruitment such as boosting social media posts through paid advertising and collaboration with the Florida State University, Network for Clinical Research, Training, and Community Engagement Service. Additionally, future work will be conducted with participants enrolled in maternal-child home visiting programs (i.e., Healthy Start, Early Head Start) which will assist with recruitment and enrollment. The percentage of participants enrolled from those screened (21%) is consistent with the Mothers and Others study (29%, Wasser et al., 2020) and higher than the INSIGHT study (8.9%, Savage et al., 2016). Seventy-two percent of participants completed our study, which is higher than Mothers and Others at 15 months (47%, Wasser et al., 2020) and slightly lower than INSIGHT at 12 months (86%, Savage et al., 2016). Noted, our study only lasted until 4 months postpartum which is a shorter duration than other studies, but these numbers demonstrate the feasibility of the LEIFc intervention.

Participant acceptability and satisfaction with the intervention are evident in review of the qualitative data. Our study participants enjoyed having the interventionist come to their house, having an individual to discuss feeding challenges and strategies with, and the additional education offered by participating in the LEIFc intervention. Quotes from participants regarding the recognition of infant feeding cues highlight aspects of responsive feeding learned by participants. Participant feedback regarding additional needs is important to consider in the refinement of the LEIFc intervention. Most notably, the refined intervention will continue through the introduction of complementary foods. We are unable to find participant satisfaction data from other responsive feeding interventions thus, unable to make comparisons.

It is important to examine differences in participants who completed all study visits and those who did not. Though it did not reach significance, participants who identified as Hispanic were less likely to complete this study. Recognizing that cultural beliefs and practices are important to interventions targeting infant feeding (de Diego-Cordero et al., 2021; Woo Baidal et al., 2015), a bilingual research assistant (RA) was added to the team, all materials were translated into Spanish and the RA was available for translation between interventionist and participant during study visits; however, no Spanish-speaking participants were enrolled. This option became available after initial recruitment and enrollment began. Though we did not ask participants’ preferred language upon enrollment, it is plausible that a greater number of Hispanic participants may have been retained if a bilingual RA was available at study initiation. Hispanic infants are at risk for overfeeding, rapid weight gain, and obesity later in life, thus, an important population to include in studies focused on the responsive infant feeding approach (Beck et al., 2018; Cartagena et al., 2015; Cartagena et al., 2014; Hu & Staiano, 2022; Stierman et al., 2021).

Although not the primary focus of this proof-of-concept study, statistical significance was examined. A significant increase in awareness of cues was seen pre-post intervention in this study. This demonstrates that participants were more aware of their infant cues and more knowledgeable in how to respond to the cues after participating in the LEIFc intervention.

Though it did not reach significance, the decrease in using food to calm the infant is also suggestive of responsive feeding by participants better understanding of their infants’ cues and knowing differences between hunger cues and cues for other needs. These results are similar to those found in prior studies that demonstrated more responsive feeding practices in participants who received a responsive parenting intervention (Daniels et al., 2013; Savage et al., 2018; Thompson et al., 2021). Though, our study did not include a control group, it is important to note that our results are from when the infant is still young, 4 months, opposed to the other studies that found this association as the child was older, 1-2 years. This early increase in responsive feeding behaviors is important to consider during this early phase of rapid growth and when the infant transitions from milk-based feedings to solid foods.

A significant increase in feeding on a schedule was seen pre-post intervention. Although this practice is not consistent with the responsive feeding approach, it is noted that many mothers return to work around 3 months postpartum and/or have others (i.e., partner, family, friends, daycare) care for their infant for an extended period of time (Bureau of Labor Statistics, 2024; Van Niel et al., 2020). Due to this transition, many mothers may have felt that a set feeding schedule was easier for another caregiver and thus, actually worked toward a schedule regardless of the mothers’ awareness of cues. Prior studies have found a risk for rapid infant weight gain in mothers who returned to work less than 12 weeks postpartum (Eagleton et al., 2019) and it is well documented that shorter durations of breastfeeding are associated with earlier return to work (Skafida, 2012; Thulier & Mercer, 2009), thus return to employment is an important variable to consider in relation to responsive infant feeding. In a larger sample size, weeks since birth when the mother returns to work should be considered as a covariate to evaluate the association with responsive infant feeding. It should also be noted that in this study, the mean scores for feeding on a schedule were low (1.38 pre and 1.64 post) and a small percentage of participants reported feeding on a schedule “often” or “always” which suggests that this may be a practice by only a small number of participants in this sample. Understanding what is different about these participants warrants further investigation.

### Strengths and Limitations

Strengths of this study include the use of a validated individualized coaching approach, SS-OO-PP-RR. SS-OO-PP-RR easily adapted from use in early language development to a responsive infant feeding intervention. An experienced interventionist implemented the intervention guided by this approach with a mean fidelity of 82. Also novel to this intervention, opposed to other responsive feeding interventions, was measurement of responsive infant feeding pre-post the intervention. This paper only reports subjective measurement but objective measurement was also preformed and will be presented at a later date. Additionally, our sample was racially diverse providing a distinction from prior responsive feeding interventions. Finally, collecting qualitative data from participants provided a perspective regarding their experience with the study and input into how to refine the LEIFc intervention moving forward.

Despite the strengths of the study, there are several limitations to note. First, the small sample size limits generalizability and the ability to analyze potential covariates. The lack of a control group prevented us from being able to determine if the increase in the responsive infant feeding approach was due to the intervention or a natural progression seen as an infant ages. We also recognize that despite an initial study visit prenatally in which education on infant feeding and responsive feeding was provided, the intervention using SS-OO-PP-RR did not occur until the infant was 2 months of age. This was potentially too late as some feeding practices were likely established by that time. Also, having only two coaching visits and one follow-up visit, the opportunity to evaluate parent-implemented intervention strategies was limited due to the number of coaching visits. Future iterations of the protocol will include a measurement of responsive infant feeding beliefs prenatally, an earlier intervention time point after birth (ideally 2 weeks postpartum), and more than two intervention visits postnatally.

Additionally, the intervention will continue through the introduction of complementary foods. Another limitation with the protocol was timing the study visit when the infant would be hungry as observation and interaction during an infant feeding session is necessary for the SS-OO-PP-RR coaching approach. Coordination of research personnel along with mother-infant schedules created challenges for all; at times the visit was rushed because the infant was very hungry when the research team arrived. This is also counterintuitive to supporting responsive infant feeding as feeding on a schedule is discouraged.

### Conclusion

This proof-of-concept study of the LEIFc intervention demonstrated that the intervention was effective, feasible to implement, and acceptable to participants. The study also provided valuable information regarding how to refine the intervention, consistent with the ORBIT framework, to move forward with behavioral intervention development. Results were consistent with prior literature on responsive infant feeding showing that the intervention decreased the use of food to calm. This study contributes to existing literature by examining the use of the responsive feeding approach pre-post the intervention, studying a racially diverse sample of mother-infant dyads, and providing qualitative data from participants on their perspective of the study. Thus, our team will move forward with refinement of the intervention and testing in a sample at the greatest risk for poor infant outcomes such as rapid weight gain and childhood obesity.

## Data Availability

All data produced in the present study are available upon reasonable request to the authors. A Data Use Agreement (DUA) may be required.

